# The quality of data-driven hypotheses generated by inexperienced clinical researchers: A case study

**DOI:** 10.1101/2024.08.12.24311877

**Authors:** Mytchell A. Ernst, Brooke N. Draghi, James J. Cimino, Vimla L. Patel, Yuchun Zhou, Jay H. Shubrook, Sonsoles De Lacalle, Aneesa Weaver, Chang Liu, Xia Jing

## Abstract

**Objectives:** We invited inexperienced clinical researchers to analyze coded health datasets and develop hypotheses. We recorded and analyzed their hypothesis generation process. All the hypotheses generated in the process were rated by the same group of seven experts by using the same metrics. This case study examines the higher quality (i.e., higher ratings) and lower quality of hypotheses and participants who generated them. We characterized the contextual factors associated with the quality of hypotheses.

**Methods:** All participants (i.e., clinical researchers) completed a 2-hour study session to analyze data and generate scientific hypotheses using the think-aloud method. Participants’ screen activity and audio were recorded and transcribed. These transcriptions were used to measure the time used to generate each hypothesis and to code cognitive events (i.e., cognitive activities used when generating hypotheses, for example, “Seeking for Connection” describes an attempt to draw connections between data points). The hypothesis ratings by the expert panel were used as the quality of the hypotheses during the analysis. We analyzed the factors associated with (1) the five highest and (2) five lowest rated hypotheses and (3) the participants who generated them, including the number of hypotheses per participant, the validity of those hypotheses, the number of cognitive events used for each hypothesis, as well as the participant’s research experience and basic demographics.

**Results:** Participants who generated the five highest-rated hypotheses used similar lengths of time (difference 3:03), whereas those who generated the five lowest-rated hypotheses used more varying lengths of time (difference 7:13). Participants who generated the five highest-rated hypotheses also utilized slightly fewer cognitive events on average compared to the five lowest-rated hypotheses (4 per hypothesis vs. 4.8 per hypothesis). When we examine the participants (who generated the five highest and five lowest hypotheses) and their total hypotheses generated during the 2-hour study sessions, the participants with the five highest-rated hypotheses again had a shorter range of time per hypothesis on average (0:03:34 vs. 0:07:17). They (with the five highest ratings) used fewer cognitive events per hypothesis (3.498 vs. 4.626). They (with the five highest ratings) also had a higher percentage of valid rate (75.51% vs. 63.63%) and generally had more experience with clinical research.

**Conclusion:** The quality of the hypotheses was shown to be associated with the time taken to generate them, where too long or too short time to generate hypotheses appears to be negatively associated with the hypotheses’ quality ratings. Also, having more experience seems to positively correlate with higher ratings of hypotheses and higher valid rates. Validity is a quality dimension used by the expert panel during rating. However, we acknowledge that our results are anecdotal. The effect may not be simply linear, and future research is necessary. These results underscore the multi-factor nature of hypothesis generation.

## Introduction

Hypothesis generation is an important and valuable process in nearly every research project. A solid hypothesis is critical and a foundation for conducting impactful research. The hypothesis marks a project’s starting line. Exploring and understanding these processes and their influencing factors are crucial for improving hypothesis generation, especially to initiate a research project. Past research into the mechanisms of hypothesis generation has focused on clinical care and pure scientific settings [1–10]. Additionally, two textbooks offer systematic introductions to scientific thinking, including hypothesis generation [11–12]. However, more insights are needed regarding the mechanisms and processes of hypothesis generation for clinical research projects, as much is still unknown regarding factors influencing hypothesis generation in such a context.

There are at least two types of research hypotheses: (1) hypotheses based on experimental observations, where researchers may observe phenomena and seek to understand them further, and (2) data-driven scientific hypothesis generation, where researchers analyze various data sets and may identify patterns or differences that prompt further exploration [13]. While both types are valuable, this case study primarily focuses on data-driven hypothesis generation.

Our team conducted a human subject study to explore hypothesis generation by clinical researchers and documented the data-driven hypothesis generation processes, i.e., analyzing data sets and developing hypotheses. We recorded their thought processes (including multiple cognitive events) via think-aloud protocol, the time it took to generate hypotheses, etc. [13–17]. The quality of the hypotheses generated in the experiment was rated by the same expert panel by using the same instrument. In this manuscript, we only focus on inexperienced clinical researchers (based on their years of experience, publications, and roles in research projects).

Patel’s studies demonstrate differences in hypothesis generation in clinical diagnosis among clinicians with different experience levels. We, therefore, separate participants based on experience levels. The study aimed to better understand the scientific hypothesis generation process, especially within a clinical research context, to provide insights into ways to facilitate and improve the process. Understanding hypothesis generation among inexperienced clinical researchers gives us insight into ways in which the process of generating hypotheses can be improved, allows for a better understanding of how outside tools and factors influence hypothesis generation and provides opportunities to improve the process at an early stage.

Additionally, our team created VIADS (a visual interactive tool for filtering and summarizing large health datasets coded with hierarchical terminologies) as a secondary data analysis tool to organize, filter, summarize, and visualize large datasets and facilitate data-driven hypothesis generation [14]. We then conducted utility and usability studies of VIADS. We have published various aspects of this research, including the study protocol for data-driven hypothesis generation among clinical researchers [14], the results of a usability study of VIADS [15], clinical research hypothesis quality assessment instruments [16], cognitive events (i.e., cognitive activities used to generate hypotheses, for example, “Seeking for Connection” describes an attempt to draw connections between data points) used during hypothesis generation between users of VIADS and those who used other tools [13], and the results of comparing hypothesis generation between two groups: VIADS users and users of other tools [17, 18].

To further explore our data, in this manuscript, we concentrated on the highest and lowest-rated quality of hypotheses and analyzed associated factors to understand why those hypotheses were rated exceptionally high or low. Subsequently, we utilized data from nine participants who generated these highest and lowest-rated hypotheses and compared their overall hypothesis generation processes and results. While other published results from the main study were based on comparisons and analyses at the group level, this case study focuses on comparisons of the individual hypotheses and participants. In this case study we aim to provide additional insights into understanding factors that influence the scientific hypothesis generation process in a clinical research context.

## Methods

### Case Study Question, Purpose, and Analysis

We aim to examine and compare the quality ratings of hypotheses and identify factors that could contribute to “higher-rated” hypotheses versus the lower-rated ones. We analyzed the time needed to create hypotheses, the number of hypotheses per participant, the valid rate of those hypotheses, and the number of cognitive events used for each hypothesis. Additionally, we considered the participants’ clinical research experience and demographics.

The question explored in this case study is: What factors may be associated with “higher-rated” hypotheses? We compared (1) the five highest-rated hypotheses (Top 5) and five lowest ones (Bottom 5) and (2) all hypotheses generated by the participants who generated the Top 5 and the Bottom 5 hypotheses.

### Background Introduction to Study Design

In the original [14, 17, 19] study, participants used identical health data sets coded with ICD9 (International Classification of Diseases–9^th^ Revision) codes, adhered to the same study scripts and think-aloud protocols (i.e., verbally “work through” and articulate what they are doing while doing it), were facilitated by the same study coordinator, and used 2 hours to generate their hypotheses. Participants were randomly assigned to two groups [14]: one utilized VIADS, and the other employed any other data analytic tool of their choice (e.g., R, SPSS, Excel). After the study session, participants completed surveys regarding their background and experience in clinical research [14]. The participants’ screen activities and audio were recorded during the study sessions.

Next, the study recordings were transcribed and coded. Initially, the time required to generate each hypothesis was examined and averaged for each hypothesis and each participant. Subsequently, cognitive events were coded using Atlas.ti 9 (a qualitative data analysis tool) for all hypotheses. For instance, the code (i.e., cognitive event) “analyze data” was assigned whenever a participant examined the data to comprehend them before formulating a hypothesis [13]. In other words, cognitive events are granular units of thought processes used by the participants while generating their hypotheses. Codes (such as “analyze data”) were used to represent cognitive events during data analysis. Two research assistants independently conducted time and cognitive event coding initially, with results being compared and consolidated. Any discrepancies were resolved by including a third team member and refining coding principles. The total number of cognitive events (codes) per participant and the average number of codes used per hypothesis were calculated.

Subsequently, an expert panel comprising seven members rated the hypotheses based on established evaluation metrics and instruments [16]. These metrics encompassed ten dimensions: novelty, clinical relevance, validity, feasibility, significance, potential benefits and risks, clarity, testability, ethicality, and interestingness. Each dimension included subitems evaluating specific aspects within the dimension (e.g., validity included clinical validity and scientific validity) [16]. Ratings were conducted using a 5-point Likert scale, and data were collected and averaged to determine the quality rating of each hypothesis [16]. During the evaluation, the expert panel utilized a three-dimensional instrument (i.e., a brief version of the instrument encompassing validity, significance, and feasibility) following a reliability check. Therefore, each hypothesis received a quality rating between 3 and 15. This case study centers on the five highest and five lowest-rated hypotheses and the participants who generated them.

This study was approved by the Institutional Review Boards of Clemson University (South Carolina, IRB2020-056) and Ohio University (18-X-192).

## Results

We organized the results into two main sections: individual hypotheses and all hypotheses generated by each participant. Each section is split into subsections comparing the highest and lowest hypotheses while examining factors that may be correlated with the ratings. Each section examines cognitive event (code) usage, time allocation, and other pertinent factors. Figure 1 provides a visual overview of the results.

**Figure 1:**
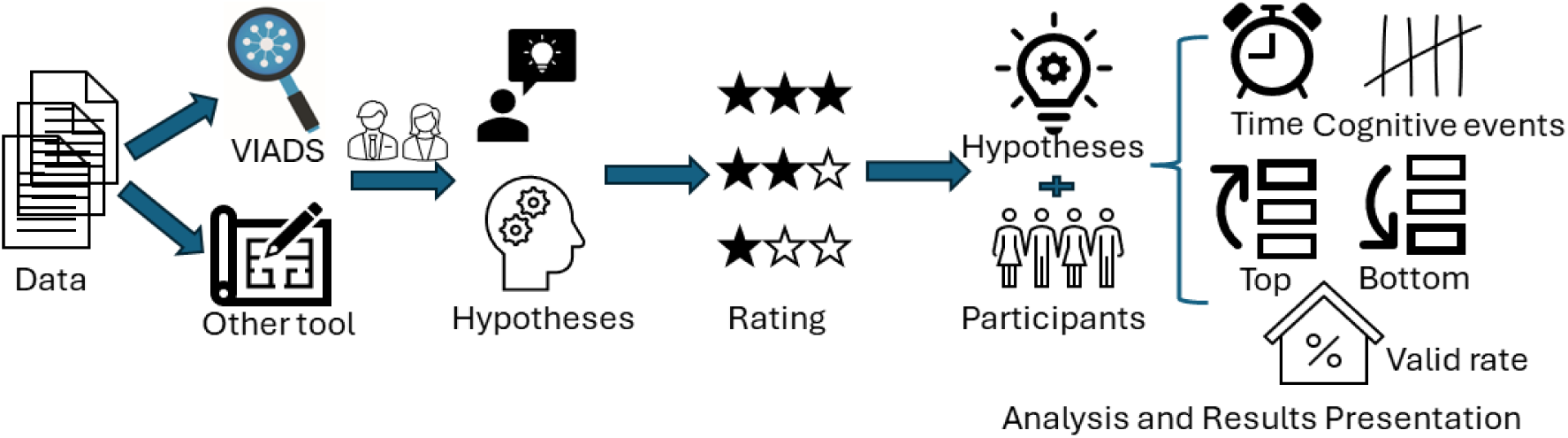
The brief study flow and the results overview *(The results are organized into two main sections: (1) specific individual hypotheses with the five highest and five lowest ratings, and (2) all the hypotheses generated by these participants from section 1.)*

### Individual Hypothesis

#### The Top 5 and the Bottom 5 hypotheses (Figure 2)

As depicted in Figure 2, the blue bars represent the Top 5 hypotheses, while the red bars show the Bottom 5 hypotheses. Additionally, Figure 2 provides information on the time required to generate each hypothesis and whether VIADS or other analytical tools were utilized in hypothesis generation. For instance, the highest-rated hypothesis, H9 by CP1 (coded participant 1; H9, hypothesis #9; NV-Non-VIADS participant), had a rating of 13 and took 5 minutes and 37 seconds to generate.

**Figure 2:**
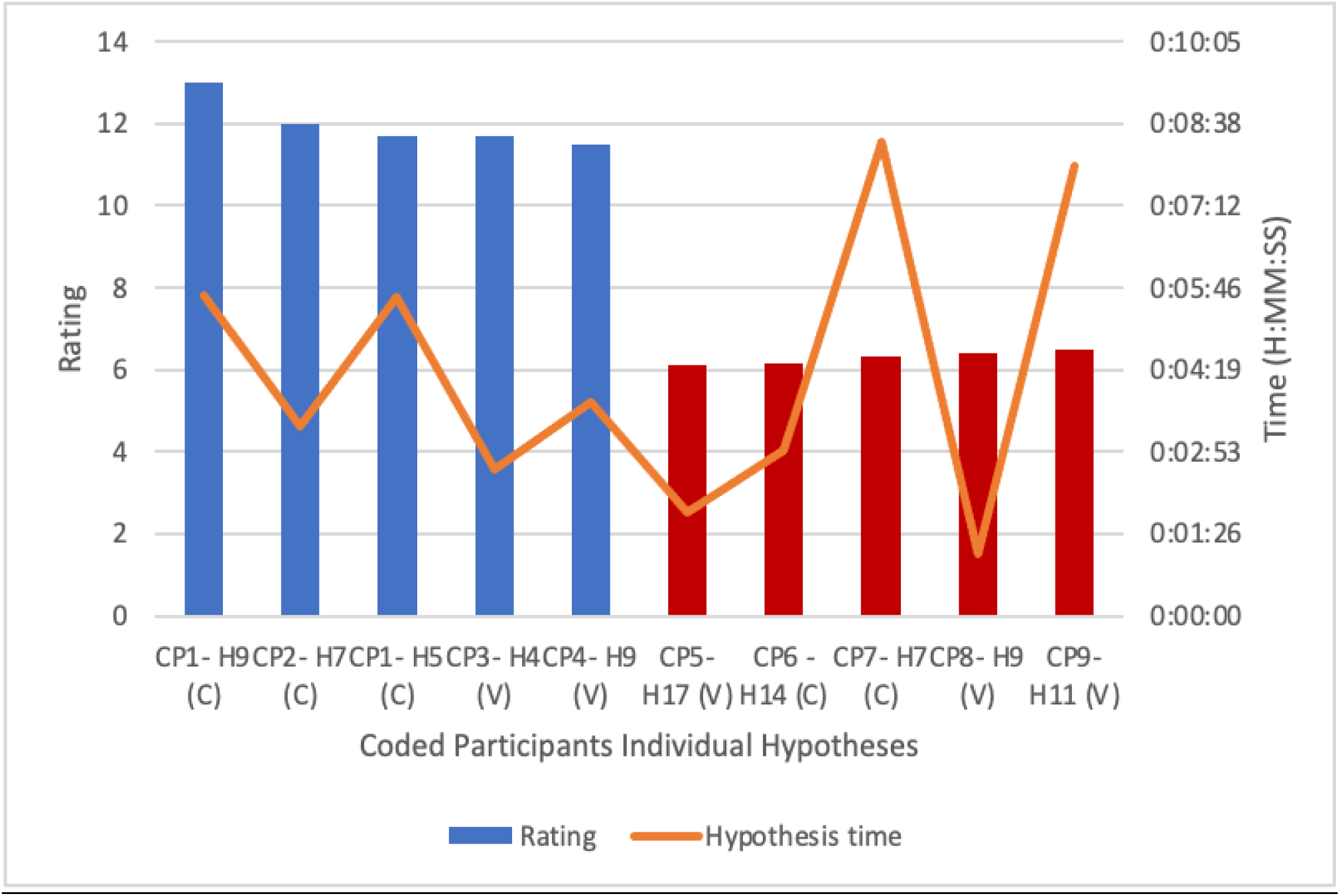
Comparison of the Top 5 (blue) and the Bottom 5 (red) Hypotheses and Time Requirements (orange). *(C-Control, V-VIADS; CP, coded inexperienced participants; H, hypothesis)*

#### Observed Differences

##### Time Usage

As illustrated in Figure 2, the time required to formulate a hypothesis varied considerably among the participants. For instance, the longest time taken to generate a hypothesis is CP7’s Hypothesis 7, which took 0:07:54 (H:MM:SS) to generate, whereas the shortest time taken to generate a hypothesis is CP8’s Hypothesis 9, which took only 0:01:05 to generate.

##### Cognitive Event Usage

As displayed in Figure 3 (below), participants generated hypotheses using a different number of cognitive events.

**Figure 3:**
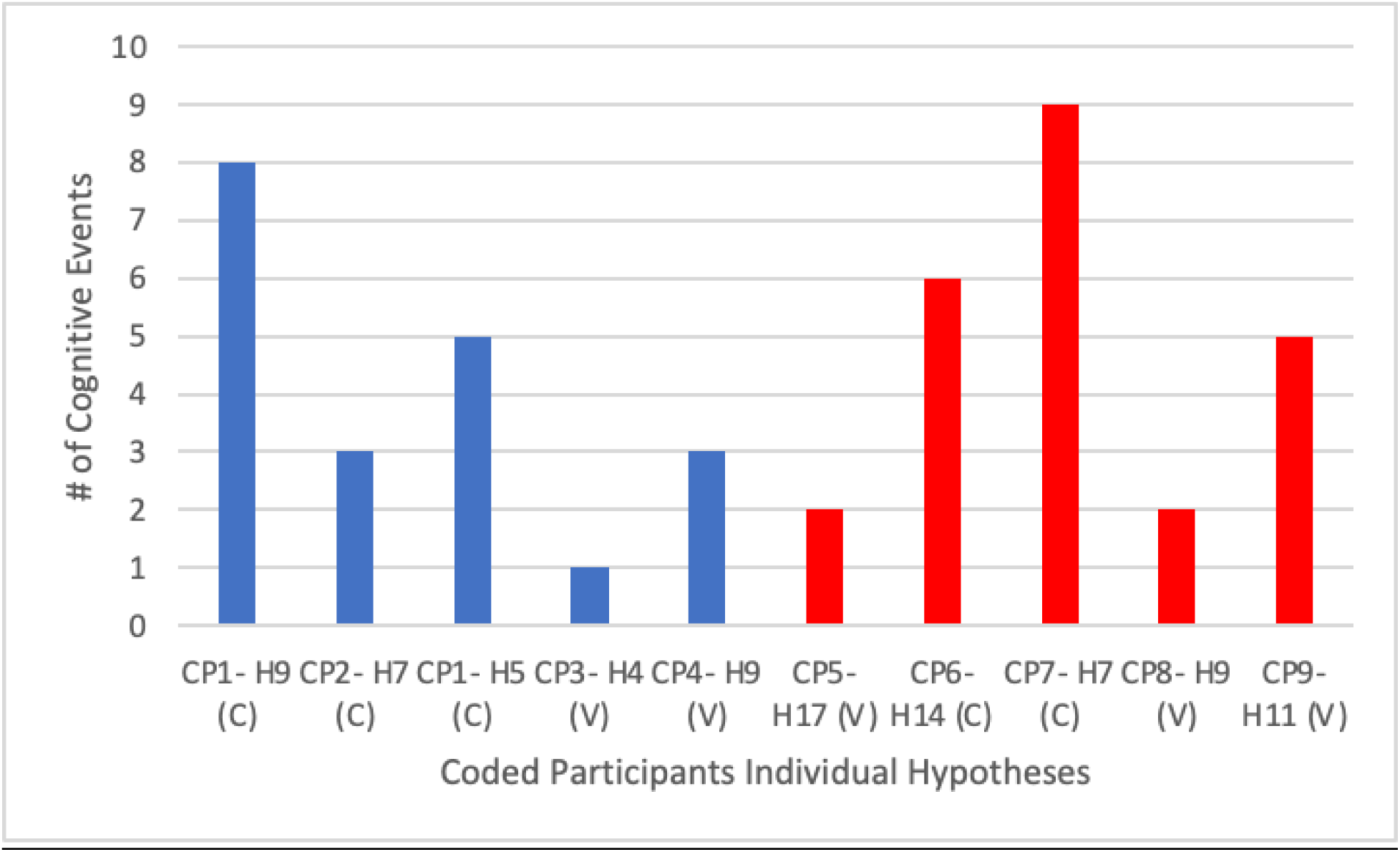
Cognitive Events Used in the Top 5 (blue) and the Bottom 5 (red) Hypotheses. (C-Control, V-VIADS; CP, coded inexperienced participants; H, hypothesis)

Figure 3 depicts the number of cognitive events utilized to generate the Top 5 and the Bottom 5 hypotheses, which exhibited considerable variability. The highest number of cognitive events in a hypothesis was Participant 7, who employed nine cognitive events in the generation of Hypothesis 7. Conversely, the lowest utilization was observed in Participant 3’s Hypothesis 4, which used just one cognitive event. Overall, participants with the Top 5 hypotheses utilized a total of 20 cognitive events to generate their hypotheses, while those with the Bottom 5 hypotheses utilized a total of 24 cognitive events.

#### Highest vs. Lowest Individual Hypothesis Comparison

Participant 1’s hypothesis 9 received a rating of 13, the highest, whereas Participant 5’s hypothesis 17 received a rating of 6.1, the lowest. Here are the two hypotheses:

- *I hypothesize that HPV vaccines made a difference in cervical cancer prevalence. To look into the last 10 to 15 years [or it depends on how long has the HPV vaccine been implemented broadly] of cervical cancer incidence data longitudinally to check if there is a correlation between the decreased cancer incidence and the implementation of the HPV vaccine broadly. (*Participant 1’s hypothesis 9)
- *#366 (ICD9 code): Cataracts had a diagnosis frequency of 746 in 2015 and 331 in 2005. The hypothesis is that more patients, especially lower economic status patients, were diagnosed early in 2015, contributing to higher diagnosis frequency in 2015. (*Participant 5’s hypothesis 17)

Next, we compared the highest and lowest individual hypothesis ratings (Table 1).

**Table 1:** Comparison of the Highest and Lowest Individual Hypothesis Ratings.

| Coded Participant | Hypothesis Rating | Average Rating/Hypothesis | Hypothesis Time | Avg. Time/hypothesis | # of valid hypotheses/total # of hypotheses | # of cognitive events | Average cognitive events/hypothesis |
| --- | --- | --- | --- | --- | --- | --- | --- |
| CP1- H9 | 13 | 10.25 | 0:05:37 | 0:05:16 | 12/15 | 8 | 5.93 |
| CP5- H17 | 6.1 | 8.819 | 0:01:49 | 0:02:09 | 19/21 | 2 | 3.14 |

Participant 1’s 9th hypothesis (highest rating) required significantly more time (0:05:37) and cognitive events (8) than Coded Participant 5’s 17th hypothesis (lowest rating), which took 0:01:49 and two cognitive events. Although both participants exhibited similar average times per hypothesis throughout the study session, Participant 1 appeared to utilize significantly more cognitive events to generate Hypothesis 9 (8) than the average cognitive events used per hypothesis (5.93). In contrast, Participant 5 seemed to employ slightly fewer cognitive events for Hypothesis 17 (2) than the average (3.14).

For comparison, Participant 1 used the following cognitive events and flow to generate hypothesis 9 (the highest): Pause/think → Analyze Data (x2) → Use Suggestion from Colleague →Analyze Data →Use Analysis Results & Seeking for Connection → Search/ask for more evidence →Use Analysis Results & Seek for Connection.

On the other hand, Participant 5 used Analyze Data →and Use Analysis Results & Seeking for Connection to generate hypothesis 17 (the lowest).

### Individual Participants

#### Highest 4 vs. Lowest 5 Participants’ All Hypotheses

Four participants generated the Top 5 hypotheses, and five participants generated the Bottom 5 hypotheses. Figure 4 shows the average quality rating per participant and the average time taken per hypothesis by each participant. For instance, CP1 (coded participant 1, NV - Non-VIADS participant), who generated the highest-rated hypothesis, had an average rating of 10.25 and took an average time of 5 minutes and 16 seconds to generate a hypothesis. In contrast, CP5 (coded participant 5, V - VIADS participant), who generated the lowest-rated hypothesis, had an average rating of 8.819 and took an average time of 2 minutes and 9 seconds per hypothesis.

**Figure 4:**
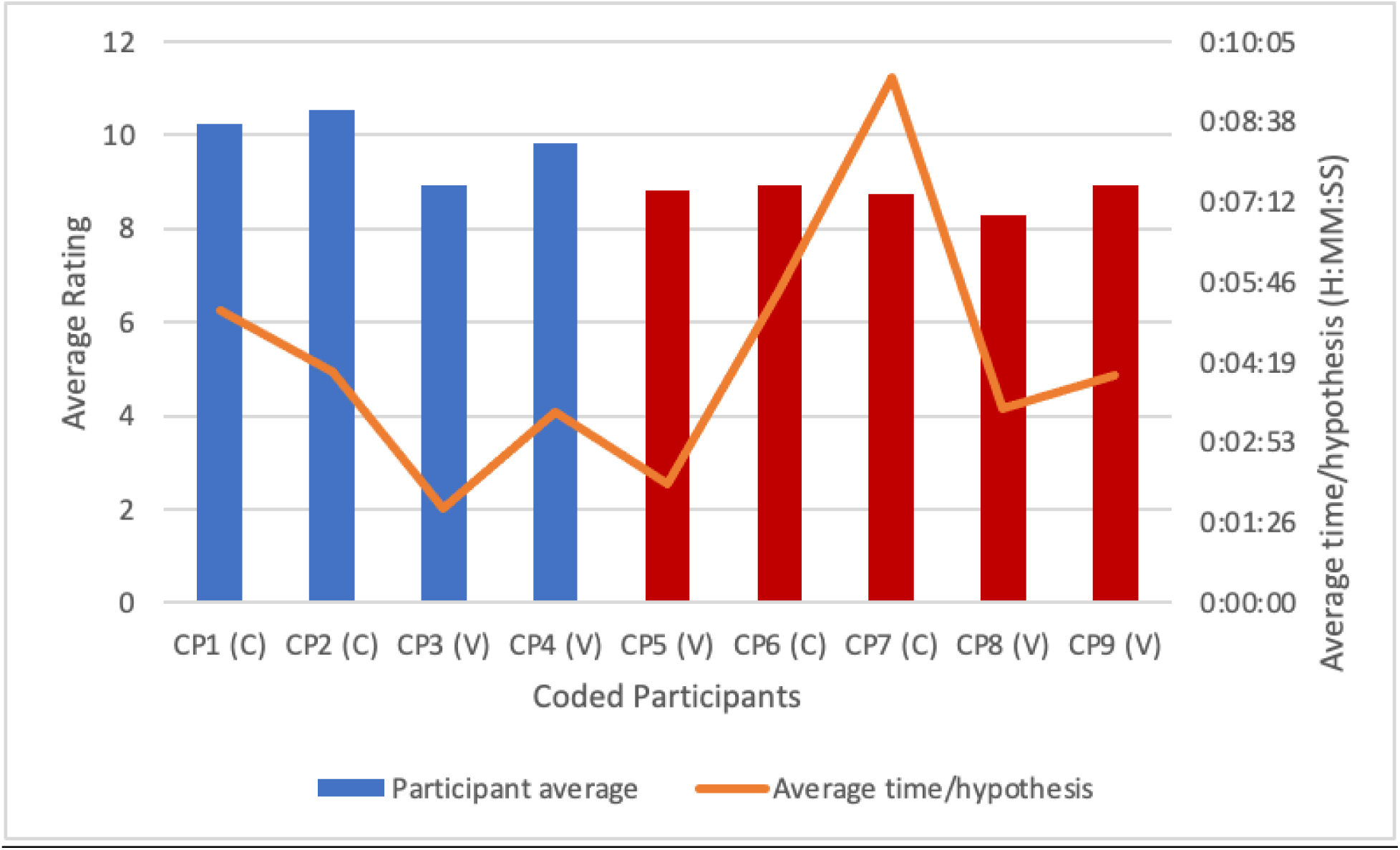
The Average Quality Rating Per Participant and Time Per Hypothesis. *Blue bars represent participants who generated the Top 5 hypotheses, while red bars denote those who generated the Bottom 5 hypotheses. (C-Control, V-VIADS; CP, coded inexperienced participants)*

#### Observed Differences

##### Time Usage

Figure 4 depicts the average time participants took to generate their hypotheses, which varied significantly. For instance, CP7 averaged the longest time, taking approximately 0:09:26 to generate each hypothesis. Conversely, CP3 averaged the shortest time, requiring approximately 0:01:42 to generate each hypothesis.

##### Cognitive Event (Code) Usage

Figure 5 shows each participant’s average cognitive events usage while generating each hypothesis.

As shown in Figure 5, CP7 used the highest average number of cognitive events per hypothesis, with an average of 6.31 codes/hypothesis. Conversely, CP3 had the lowest average number of cognitive events per hypothesis, with an average of 2.60 codes/hypothesis.

**Figure 5:**
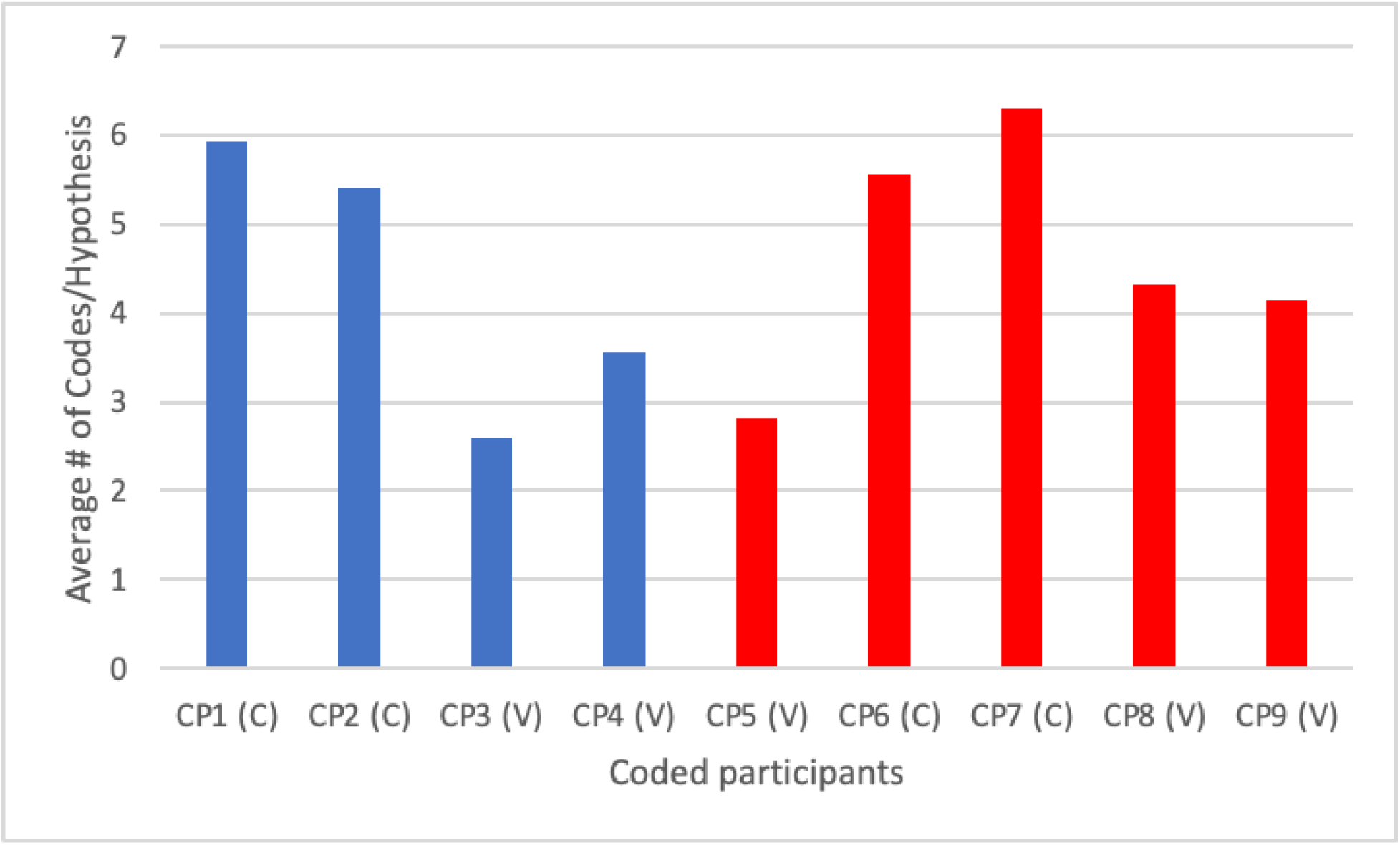
Average Number of Cognitive Events Per Hypothesis per Participant. *Blue bars represent participants who generated the Top 5 hypotheses (corresponding with Figures 2, 3, and 4), while red bars denote participants who generated the Bottom 5 hypotheses (also corresponding with Figures 2, 3, and 4). (C-Control, V-VIADS; CP, coded inexperienced participants)*

##### Valid Rates

During the hypothesis quality rating process, any hypothesis scored at “1” (the lowest rating) for validity by three or more experts was considered invalid. Among participants with the Top 5 hypotheses, CP1 had 12 valid hypotheses out of 15 (80%), CP2 had nine valid hypotheses out of 10 (90%), CP3 had eight valid hypotheses out of 15 (53.33%), and CP4 had eight valid hypotheses out of 9 (88.89%). For participants with the Bottom 5 hypotheses, CP5 had 19 valid hypotheses out of 21 (90.48%), CP6 had 13 valid hypotheses out of 16 (81.25%), both CP7 and CP8 had five valid hypotheses out of 13 each (38.46%), and CP9 had seven valid hypotheses out of 14 (50%).

##### Demographics

Demographics and experiences provide valuable insights into participants’ backgrounds, potentially influencing their approach to hypothesis generation and the subsequent ratings of hypotheses. Table 2 presents these details.

**Table 2:** Participants’ Demographics and Experiences.

| Coded Participant | YOE in Hypothesis Generation | YOE in Study Design | YOE in Data Analysis | TOR in Hypothesis Generation | TOR in Study Design | Publication # |
| --- | --- | --- | --- | --- | --- | --- |
| CP1 | >= 5 and < 10 | > 2 and < 5 | > 2 and < 5 | Collaborator role | Collaborator role | <5 |
| CP2 | > 2 and < 5 | > 2 and < 5 | <= 2 | Leading role | Leading role | <5 |
| CP3 | <= 2 | <= 2 | <= 2 | Collaborator role | Collaborator role | <5 |
| CP4 | <= 2 | <= 2 | <= 2 | Collaborator role | Collaborator role | <5 |
| CP5 | > 2 and < 5 | <= 2 | <= 2 | Leading role | Collaborator role | <5 |
| CP6 | > 2 and < 5 | > 2 and < 5 | > 2 and < 5 | Collaborator role | Collaborator role | <5 |
| CP7 | <= 2 | <= 2 | > 2 and < 5 | Collaborator role | Collaborator role | <5 |
| CP8 | <= 2 | > 2 and < 5 | <= 2 | Research<br>Coordinator | Research<br>Coordinator | <5 |
| CP9 | <= 2 | <= 2 | <= 2 | Mixed; Collaborator<br>& Leading Role | Collaborator role | <5 |

*YOE, years of experience; TOR, type of role. Participants highlighted in light blue have the highest-rated hypotheses, while those highlighted in red have the lowest.*

#### Comparing Participants with the Highest and Lowest Average Ratings

Table 3 compares the highest and lowest average hypotheses between the participants discussed in the case study (CP2; CP8), including the average rating for their hypotheses (10.55; 8.298), the average time to generate each hypothesis (0:04:10; 0:03:29), the valid rates of their hypotheses (90%; 38.46%), and the average number of cognitive events used per hypothesis (5.4; 4.31). As can be seen in Table 3, CP2 averaged a longer time per hypothesis, had a higher valid rate, and used more cognitive events per hypothesis in comparison to CP8.

**Table 3:**
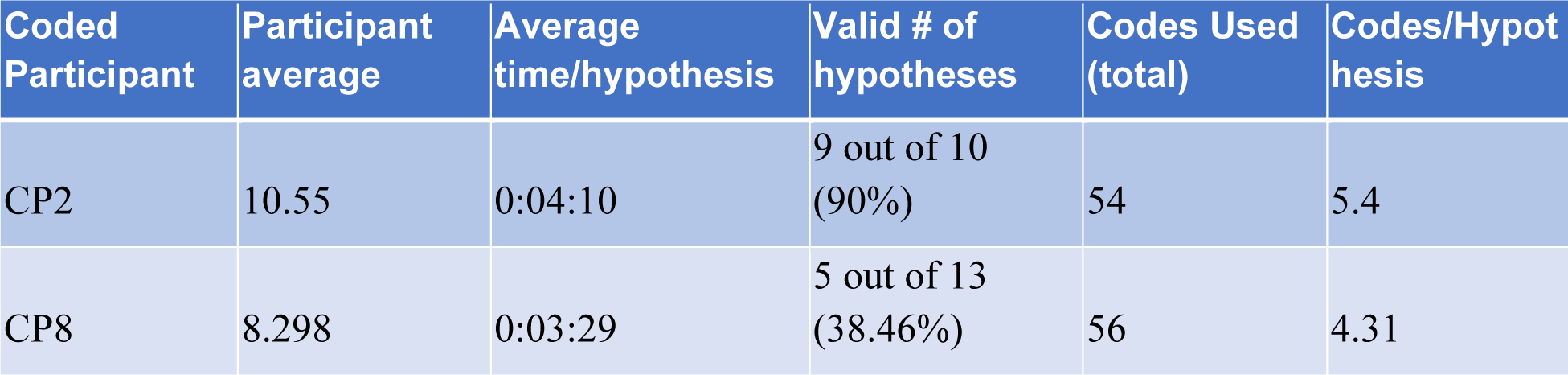
Comparison of the Highest and Lowest Average Hypothesis Ratings.

#### Summary of Results

Participants who generated the Top 5 hypotheses used similar lengths of time per hypothesis (i.e., difference 0:03:03) compared to those who generated the Bottom 5 hypotheses (i.e., difference 0:07:13). Participants who generated the Top 5 hypotheses also utilized slightly fewer cognitive events on average compared to the ones with the Bottom 5 hypotheses (4 per hypothesis vs. 4.8 per hypothesis). When comparing the two participants with the highest and lowest-rated hypotheses, the highest-rated hypothesis required more time and cognitive events than the lowest-rated hypothesis (0:05:37 vs. 0:01:49; 8 vs. 2). Comparing the participants who generated the Top 5 and the Bottom 5 hypotheses utilizing their total hypotheses generated throughout the study session, the participants with the Top 5 hypotheses again had a shorter range to generate each hypothesis on average (0:03:34 vs. 0:07:17). They used fewer cognitive events per hypothesis (3.498 vs. 4.626). They also had a higher valid rate of hypotheses (75.51% vs. 63.63%) and generally had more experience with clinical research. When comparing the two participants who generated the highest and the lowest rated hypotheses and their total hypotheses in this case study, the higher-rated participant utilized slightly longer time and more cognitive events per hypothesis (0:04:10 vs. 0:03:29; 5.4 vs. 4.31) on average and had a much higher percentage of valid hypotheses (90% vs. 38.46%).

## Discussion

### The quality of the hypotheses generated

Our purpose was to delve into the hypothesis generation process within a clinical research context, recognizing that examining individual participants and their hypotheses can yield valuable insights about how and where technology support may be valuable. As depicted in Figure 2, we compared the Top 5 to the Bottom 5 hypotheses.

First, concerning ***time usage***, a notable contrast emerges in the time taken to create these hypotheses when comparing the Top 5 to the Bottom 5. Participants in the Top 5 showed fewer variations in the time taken to generate a hypothesis (difference 0:03:03) as compared to participants in the Bottom 5 (difference 0:07:13). The latter is more than twice the former.

Another crucial factor to consider is ***cognitive events*** used in generating hypotheses. We noted the Top 5 group utilized slightly fewer cognitive events (4/hypothesis) than the Bottom 5 (4.8/hypothesis) on average, i.e., the Bottom 5 used 20% more cognitive events per hypothesis on average.

### Results Interpretation: the Highest versus the Lowest Hypothesis

Comparing the highest and lowest-rated hypotheses sheds light on the hypothesis generation. CP1’s 9th hypothesis received the highest rating among all hypotheses, scoring 13, while CP5’s 17th hypothesis received the lowest rating at 6.1. CP1 maintained an average hypothesis rating of 10.25, generating 15 hypotheses (12 of which were valid), whereas CP5 achieved an average hypothesis rating of 8.819, generating 21 hypotheses (19 of which were valid). Moreover, demographic data revealed that CP1 possessed more experience in clinical research than CP5. These differences in prior experience likely contributed to the difference in the quality ratings of their hypotheses.

Another interesting observation is the difference between time usage in generating these two hypotheses. On average, CP1 took 5 minutes and 16 seconds to create a hypothesis (the highest-rated hypothesis, H#9 took 5 minutes and 37 seconds), whereas CP5 took only 2 minutes and 9 seconds to generate a hypothesis on average (the lowest-rated hypothesis, H#17 took 1 minute and 49 seconds).

Literature indicates individuals with high time urgency tend to generate lower-rated hypotheses [20]. CP5 had to use a tool that he/she had only learned just an hour before the study session (VIADS), which could have inadvertently resulted in a feeling of urgency and cognitive overload, thereby potentially negatively affecting the quality ratings of CP5’s hypotheses. It is also plausible that cognitive overload from attempting to learn to use VIADS could have influenced these results negatively. Findings from a study by Dasgupta et al. support the idea that often, the “brain trades off accuracy and computational cost to make efficient use of its limited cognitive resources to approximate probabilistic inference” [21].

Regarding cognitive events, CP1 utilized 5.93 cognitive events/hypothesis on average. For the highest-rated hypothesis (H#9), CP1 employed eight cognitive events, whereas CP5 utilized an average of 3.14 cognitive events/hypothesis and only two for the lowest-rated hypothesis (H#17). Furthermore, other participants with higher-rated hypotheses utilized considerably fewer cognitive events, such as CP3, who used one cognitive event for H4 (rating of 11.7) and averaged 2.60 codes/hypothesis. It should be noted that this was CP3’s 4th hypothesis, and it is possible that the participant got used to the process fairly quickly, possibly resulting in the lowered usage of cognitive events. However, more data are needed to reveal the relationships between the number of cognitive events used and the quality of hypotheses generated.

### Results Interpretation: Participants Comparison

Examining the ***time usage*** among the participants who generated the Top 5 and the Bottom 5 hypotheses reveals similar differences. When examining all hypotheses generated throughout 2-hour study sessions, participants who generated the Bottom 5 hypotheses exhibited more varied time usage (difference 0:07:17) compared to the participants of the Top 5 (difference 0:03:34, Figure 4). This aligns with the findings from the comparison of individual hypotheses.

Analyzing the ***cognitive events*** utilized by these participants exhibited comparable differences. It is evident that participants of the Top 5 employed an average of 3.498 codes per hypothesis, whereas the participants of the Bottom 5 utilized an average of 4.626 codes per hypothesis. This finding partially supports the comparison between individual hypotheses of the Top 5 and the Bottom 5, suggesting that lower cognitive event usage may positively influence the ratings of generated hypotheses.

On the contrary, the hypotheses’ ***valid rate*** demonstrates a positive association with higher-rated hypotheses compared to those with lower ratings. The participants of the Top 5 generated 37 (out of 49, 75.51%) valid hypotheses, whereas participants of the Bottom 5 had 49 (out of 77, 63.63%) valid hypotheses.

Interestingly, there are notable discrepancies when examining the ***demographics and experiences*** of participants in this case study. For instance, CP1 has the most extensive experience in hypothesis generation, study design, and data analysis, aligning with the expectation that greater experience would yield higher-rated hypotheses. However, outliers such as CP3 and CP4, with minimal experience (i.e., less than two years of experience in hypothesis generation, study design, and data analysis) and collaborator roles, produced some of the highest-rated individual hypotheses. Conversely, participants like CP5, despite holding prominent roles in hypothesis generation and possessing more experience in the field, produced one of the lowest-rated individual hypotheses. The results suggest the relationships between one’s experience level and the quality of hypotheses may be more complicated.

### Results Interpretation: The Participants with the Highest and the Lowest Average Ratings

Isolating the top and bottom average hypothesis scores allows for a more nuanced examination of the quality of the hypotheses generated. CP2 attained the highest average hypothesis score among the participants, with an average score of 10.55 per hypothesis, while CP8 garnered the lowest score, with an average of 8.298. CP2 achieved both a higher score and a higher valid rate of hypotheses compared to CP8 (10.55 vs. 8.298; 9/10, i.e., 90% vs. 5/13, i.e., 38.46%), while CP2 took longer on average to generate a hypothesis and employed more cognitive events (0:04:10 vs. 0:03:29; 5.4 vs. 4.31).

Regarding demographic factors, CP2 has more experience in hypothesis generation than CP8, while both participants had similar experience levels in data analysis and study design. CP2 held a leading role, while CP8 served as a research coordinator in the past. CP2 maintained a slight advantage in experience concerning hypothesis generation, which could have influenced the rating and validity of their hypotheses.

### Strengths and Limitations

One of the strengths of this study lies in its detailed examination and comparison of hypothesis generation through different lenses: at the hypothesis level, at the participant level, as well as examining them individually or as a group. We compared the time needed and cognitive events used during hypothesis generation and the participants’ demographic factors based on hypothesis quality ratings. Currently, limited research focuses on scientific hypothesis generation, particularly in initiating research projects and understanding the cognitive processes involved. This study fills that gap by providing systematic comparisons and valuable insights with comprehensive raw data collected during the hypothesis generation process.

Another strength is the detailed analysis of a small cohort of 9 participants. While this limits the generalizability of the findings, it enables a more thorough comparison between individuals who generated higher-rated and lower-rated hypotheses. This approach offers a nuanced understanding of differences in their hypothesis generation processes and also provides an important starting point to explore the complex and crucial process. This manuscript also complements our prior reported results, which focused on group comparisons [17].

However, the study also has its limitations. First, the time constraint of the primary study session, capped at 2 hours, may have influenced participants’ ability to generate hypotheses naturally, potentially introducing bias [20]. Additionally, since participants utilized the think-aloud protocol during the study, only consciously articulated processes were captured.

Furthermore, this case study provides insights into individual hypotheses and participants, among inexperienced clinical researchers only; any comparisons involving broader participant samples should be approached with caution [17–18].

### Future Work

There are a few areas where this study could be explored. A substantial amount of data from participants with average ratings were excluded by focusing solely on participants with the highest and lowest-scoring hypotheses. Incorporating this middle-ground data could provide a more comprehensive understanding of the hypothesis generating process and potentially reveal additional insights. Furthermore, this case study exclusively examined inexperienced clinical researchers. Future research could include experienced clinical researchers. Their perspectives and approaches to hypothesis generation may offer additional valuable insights. Furthermore, additional data analysis of the think-aloud protocols may provide further insights into understanding the thought process of hypothesis generation [22, 23].

## Conclusion

Demographics and experiences emerged as influential factors, with higher levels of experience associated with generating higher-rated hypotheses. Additionally, a higher validity rate of hypotheses was correlated with producing higher-rated hypotheses.

The comparison between the highest and lowest individual hypotheses and the participants with the highest and lowest average hypotheses revealed consistent findings across various factors. Notably, outliers in hypothesis generation, characterized by either excessively short or prolonged usage of time, were negatively associated with the quality ratings of hypotheses. This trend was evident in both the individual and total hypothesis comparisons. Fewer cognitive events were used for the participants from the Top 5 group, possibly showing that they required less cognitive effort to process the data.

While this study offers valuable insights into factors affecting hypothesis quality, it is important to note that this explorational study results are from a few participants. The next potential steps in this study will include a larger sample of participants and also include experienced clinical researchers. A better understanding of the process will guide us to leverage the technology needed to facilitate an efficient and effective hypothesis generation process.

## Acknowledgment

We would like to thank all participants and expert panel members for participating in this study. Without their significant contribution, none of the work would be possible. The project was supported by a grant from the National Library of Medicine (R15LM012941) and was partially supported by the National Institute of General Medical Sciences of the National Institutes of Health (P20 GM121342). This work has also benefited from research training resources and the intellectual environment enabled by the NIH/NLM T15 SC BIDS4Health research training program (T15LM013977).

## Author contributions

XJ, YZ, VLP, and JJC originally designed the study. JHS and SDL provided significant input and feedback on the study; JJC, JHS, and SDL are instrumental in helping recruit participants. CL provided significant technical guidance and support for the study. XJ collected data; MAE and BNP analyzed data and AW helped with setting up the hypothesis evaluation; MAE prepared the first draft; XJ, VLP, JJC, and YZ provided substantial revisions. All co-authors revised the manuscript significantly and agreed on the final version.

## Data availability

More detailed, analyzed, and organized data are available upon request to the corresponding author; requests for the raw data are considered on a case-by-case basis.

